# Cost-Aware Active Feature Acquisition for Differential Diagnosis under Realistic Clinical Availability Constraints

**DOI:** 10.64898/2026.08.30.26361745

**Authors:** Joseph Bingham, Netanel Arussy

## Abstract

Active Feature Acquisition (AFA) adaptively selects which diagnostic test to order next and offers a route to reduce unnecessary laboratory testing in acute care. Existing clinical AFA evaluations, however, assume every feature can be retrieved on demand and split data at the visit level, both of which inflate apparent performance. We re-evaluate cost-aware AFA under constraints designed to reflect deployment. From MIMIC-IV we constructed a cohort of 64,766 acute admissions (39,884 patients; 21 conditions; 55 features in 30 test panels) with a patient-level split, a 12-hour decision cutoff, and a per-patient availability mask from what was actually measured, and priced panels using the 2026 Medicare fee schedule under panel-level billing. We evaluated EIG-Cost, which scores each panel by Monte-Carlo Expected Information Gain penalised by its dollar cost, against eight published methods across budgets $30–$60 over five patient-level resamples. At a $30 budget, EIG-Cost achieved the highest macro-F1 (**0.188**, 95% CI **[0.185, 0.191]**) at the lowest cost ($17.28), exceeding the strongest baseline in all five resamples (***p* < 0.001**; Cohen’s ***d* = 4.0**), and led at every budget. Three of the eight methods collapsed to a vitals-only baseline (macro-F1 **≈ 0.040**), acquiring nothing even at higher budgets, a genuine failure to adapt to availability rather than a budget limitation. Despite modest absolute accuracy, EIG-Cost’s probabilities were well-calibrated (expected calibration error **0.048**). Under realistic availability constraints, clinical AFA is substantially harder than full-availability benchmarks imply, several published methods fail outright, and cost-aware information-gain scoring is a robust choice in this harder setting.

## 1 Introduction

Acute hospital care frequently presents clinicians with a differential diagnosis requiring targeted laboratory investigation. Laboratory overutilisation is well documented: an estimated 20–30% of inpatient laboratory orders provide no incremental diagnostic value van Walraven and Naylor (1998), and the Choosing Wisely initiative cho (2012) has catalogued hundreds of tests frequently ordered without clear indication. Deciding *which* test to order next, given what is already known about a patient, is therefore a consequential and recurring problem.

Active Feature Acquisition (AFA) formalises this decision as sequential machine learning Settles (2009): an agent observes a partial feature vector, maintains a predictive model over diagnoses, and selects the next feature to acquire, trading information against cost. Recent methods use deep generative models Ma et al (2019); Gadgil et al (2024) or reinforcement learning Janisch et al (2019); Shim et al (2018) to learn acquisition policies.

Despite this progress, clinical AFA has largely been evaluated under assumptions that do not hold at the bedside, and two are particularly consequential. *First*, most evaluations assume **full availability**: the agent may acquire any feature at any time, as though every laboratory value could be retrieved on demand. In reality, at an early decision point most values have not been measured and cannot be produced retroactively. The relevant question is which of the *available* tests to order, not which of all conceivable tests. *Second*, evaluations commonly use **visit-level splits** that place different admissions of the same patient on both sides of the train/test boundary, leaking patient-specific signal and inflating apparent performance. A third, more technical gap is that most methods express acquisition budgets as feature *counts* rather than money, ignoring the large variation in test cost (from $0 for vital signs to tens of dollars for specialised assays) and precluding direct optimisation of cost-efficiency.

We revisit cost-aware AFA after removing these assumptions. We construct a MIMIC-IV cohort with a strict patient-level split, a fixed 12-hour decision-time cutoff, and a per-patient *availability mask* that restricts acquisition to features actually measured for that patient within the window. We price panels with a clinically faithful billing model in which ordering a panel charges its cost once and returns all of its components. Under these constraints we evaluate EIG-Cost (Expected Information Gain penalised by dollar cost) against eight published methods.

This paper makes three contributions. **(1)** A realistic-constraint evaluation protocol for clinical AFA, combining patient-level splitting, a decision-time cutoff, empirical per-patient availability, and panel-level dollar costing. **(2)** Evidence that, under these constraints, several published AFA methods collapse to a vitals-only baseline and do not recover at higher budgets, in contrast to their apparent competitiveness under full-availability assumptions. **(3)** Evidence that cost-aware information-gain scoring (EIG-Cost) is Pareto-dominant on the cost–performance frontier in this harder setting, with its advantage concentrated at the tight budgets most relevant to practice. Figure 1 summarises the method and the headline result.

**Fig. 1.**
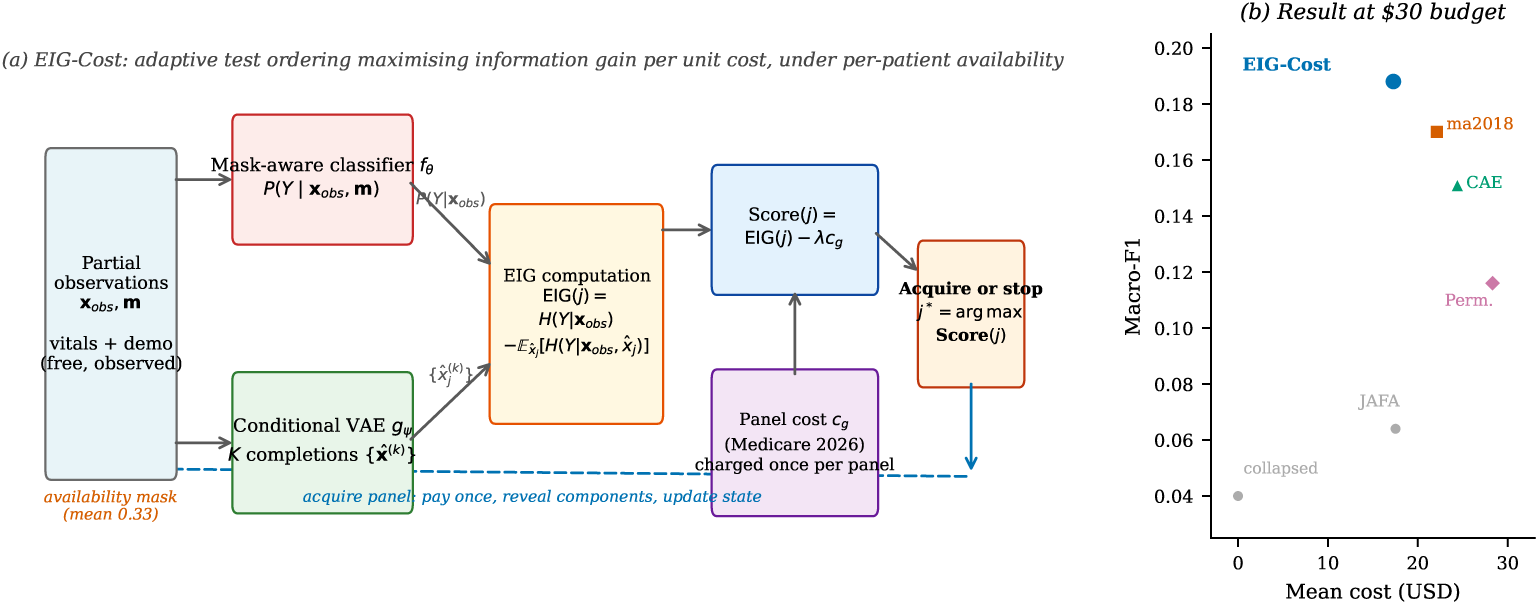
Overview. (a) At each step, EIG-Cost scores every unacquired, *available* test panel by its expected reduction in diagnostic uncertainty (estimated with a mask-aware classifier and a conditional generative model) minus its dollar cost, acquires the best panel if its score is positive and affordable (charging the panel cost once and revealing its components measured for that patient) and otherwise stops and predicts. (b) Under realistic availability constraints on a MIMIC-IV cohort, EIG-Cost achieves the best macro-F1 at the lowest realised cost at a $30 budget, while three of eight methods collapse to a vitals-only floor. All values trace to the verified five-seed results reported in Section 3.

## 2 Methods

### 2.1 Cohort construction

We used MIMIC-IV v2.2 Johnson et al (2023) (Beth Israel Deaconess Medical Center). We identified adult acute admissions associated with at least one of 21 acute diagnostic conditions, ascertained from ICD-9/10 discharge codes; when an admission matched multiple conditions, the primary discharge diagnosis was assigned as the label. The resulting cohort comprises 64,766 admissions from 39,884 unique patients. Each admission is characterised by 55 laboratory and vital-sign features grouped into 30 test panels.

#### Decision-time cutoff

To reflect an early diagnostic decision point, each admission is anchored 12 hours after arrival, and only information that would be available by that time is used. This avoids the label leakage that arises when values recorded near discharge are treated as available at admission.

#### Patient-level split

We partitioned the cohort into training (41,649 admissions), validation (10,233), and test (12,884) sets at the *patient* level, so that no patient contributes admissions to more than one partition. Zero subject overlap across partitions was verified directly from the data.

#### Availability

For each admission we constructed a binary availability mask indicating which of the 55 features were actually measured within the 12-hour window.

Mean availability across the test set is 0.330; eight features (demographics and vital signs) are available for all patients at zero cost, while several specialised assays are available for only a small fraction. Acquisition is restricted to available features: the agent cannot order a value that was never collected for that patient (Figure 2).

**Fig. 2.**
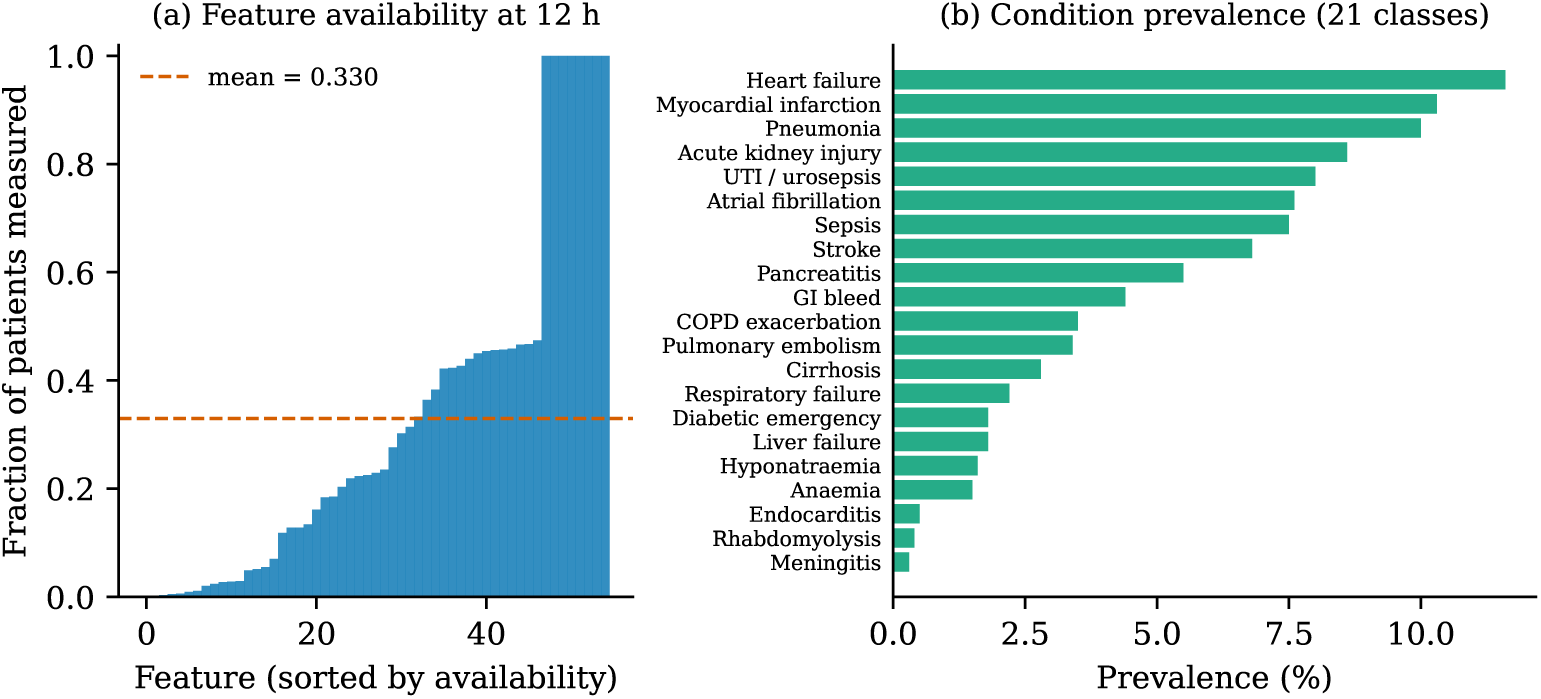
Cohort characteristics. (a) Per-patient feature availability within the 12-hour decision window, sorted; eight demographic and vital-sign features are available for all patients, while many laboratory panels are measured for only a minority (mean availability 0.330). (b) Prevalence of the 21 diagnostic conditions, ranging from heart failure (11.6%) to meningitis (0.3%), motivating macro-F1 as the headline metric.

#### Costs

Each panel was assigned its 2026 Medicare Physician Fee Schedule reimbursement Centers for Medicare & Medicaid Services (2025). We use a panel-level billing model: the first component acquired from a panel charges the panel’s list cost once, and any further components of that same panel are then free. Acquisition reveals only the components that were actually measured for the patient within the 12-hour window (see availability, above); a panel is never charged for values that were not collected. The full-workup cost across all 30 panels is $408.80 (Table 1). Demographics and vital signs are pre-acquired at zero cost.

**Table 1.**
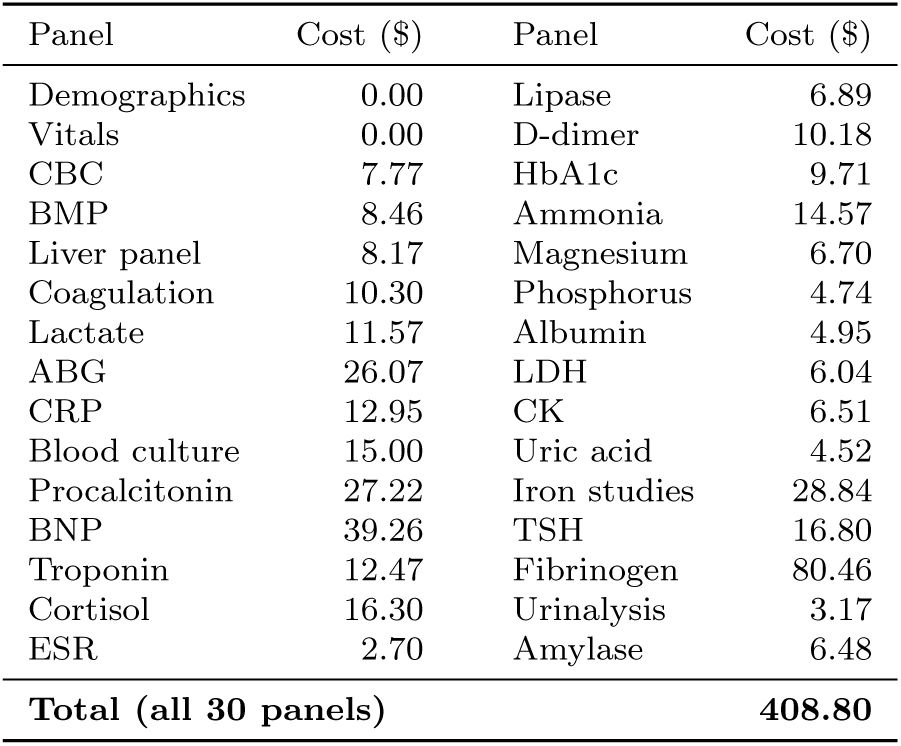
Acquisition cost of each test panel (2026 Medicare Physician Fee Schedule). Demographics and vital signs are pre-acquired at zero cost; the full-workup cost across all 30 panels is $408.80. Under panel-level billing, a panel is charged once and all of its component features are then revealed.

#### Missing data

Unmeasured features are structurally missing rather than missing at random: at the decision point they were simply not ordered. We represent this explicitly with the availability mask and a mask-aware classifier (below) that receives both the observed values and an indicator of which features are observed, so that “not measured” is distinguished from “measured as zero” without imputation.

### 2.2 The EIG-Cost method

#### Problem formulation

Let **x** *∈* ℝ*^d^* be a patient’s complete feature vector and *Y* ∈ {1,…, *K*} the diagnostic label. A binary mask **m** ∈ {0, 1*}^d^* indicates acquired features and **x**_obs_ = **x** ⊙ **m** is the partial observation. Features are grouped into *G* = 30 panels; acquiring panel *g* reveals its features at cost *c_g_ ≥* 0 (USD), charged once per panel. The agent selects available panels sequentially until cumulative cost exceeds a budget *B* or it stops.

#### Mask-aware classifier

Classifiers trained on complete data are unreliable under partial observation. We train a classifier *f_θ_* on the concatenated input [**x**_obs_; **m**] ∈ ℝ^2^*^d^*,

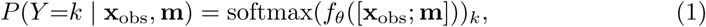

so it distinguishes observed zeros from missing entries without imputation. Each feature is independently masked with probability 0.7 during training, exposing the model to partial-observation distributions. The network is a three-layer MLP ([256, 128, 64], batch normalisation, ReLU, dropout 0.2).

#### Conditional generative model

Estimating information gain requires samples from *P* (*x_j_ |* **x**_obs_, **m**). We model this with a conditional variational autoencoder Sohn et al (2015) with encoder *q_ϕ_*(**z** *|* **x**_obs_, **m**) and decoder *p_ψ_*(**x** *|* **z**, **m**), trained on the evidence lower bound

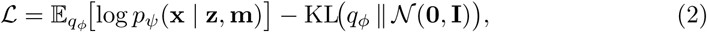

with reconstruction computed over unobserved features only. At inference, *K*=50 Monte Carlo completions are drawn per candidate feature.

Concretely, during training we sample an observation mask **m** per example (as for the classifier, each feature is independently retained with the same random masking scheme), split each example into its observed part **x**_obs_ and its held-out part **x**⊙(1−**m**), and condition the encoder and decoder on **x**_obs_ and **m**. The reconstruction term in Equation 2 is evaluated only on the held-out features **x** ⊙ (1 − **m**): the model is trained to predict what was masked given what was observed, which is exactly the conditional *P* (*x_j_ |* **x**_obs_, **m**) that the information-gain estimate requires. We do not add a separate observed-feature reconstruction term; the observed values enter only as conditioning input. At inference the same conditioning is applied with the patient’s true observed features fixed, and completions of an unobserved candidate feature are drawn from the decoder.

#### Cost-penalised scoring

With current predictive entropy H(*Y |* **x**_obs_, **m**) = Σ_k_ *P* (*Y* =*k |* **x**_obs_, **m**) log *P* (*Y* =*k |* **x**_obs_, **m**), the expected information gain of candidate feature *j* is approximated as

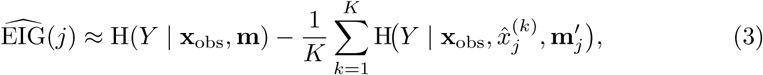

where **m**_j_*^′^* sets bit *j* to 1 and 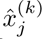 is the *k*th completion. The acquisition score subtracts the marginal dollar cost of feature *j*,

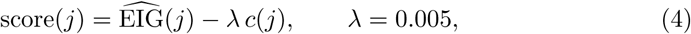

and the agent selects *j*\* = arg max*_j∈U_* score(*j*) over unacquired, *available* features *U*, stopping if the maximum score is negative or the budget is exhausted.

#### Scoring is per feature; billing is per panel

Two points are worth making explicit, as they distinguish this setting from feature-count benchmarks. First, information gain is scored at the level of the individual feature, not the panel: we do not compute a joint EIG over all components of a panel. The cost term *c*(*j*) is the *marginal* panel cost: the parent panel’s list price if none of its components has yet been acquired, and zero otherwise, so the first feature drawn from a panel bears the full panel cost and any subsequent feature from the same panel is free. This is a deliberate approximation: it keeps scoring tractable (a single feature at a time) while making the cost accounting clinically correct (a panel is billed once). It does not model intra-panel complementarity, i.e. cases where two components of a panel are jointly but not individually informative; we return to this in the limitations. Second, acquisition respects the per-patient availability mask: “acquiring” a panel reveals only those of its components that were actually measured within the 12-hour window for that patient, never values that were not collected. A feature that is unavailable for a patient is never scored or acquired for that patient, regardless of its panel’s cost. All candidate features are scored in a single batched forward pass over the *|U| × K* modified feature vectors.

#### Computational footprint

EIG-Cost runs on CPU without a GPU. On a standard server CPU, a full acquisition trajectory takes on average 180 ms per patient (median *∼* 155 ms, 95th percentile *∼* 480 ms), or about 47 ms per acquisition step, with a mean of 3.8 acquisitions per patient at the $30 budget. This is well within the latency budget of an interactive decision-support tool and does not require specialised hardware.

### 2.3 Baselines and evaluation

We compared EIG-Cost against eight published methods spanning the main AFA families: **ma2018** Ma et al (2019) (generative, EDDI-style, with the external classifier); **CAE** Covert et al (2023) (mutual-information maximisation); **DIME** Gadgil et al (2024) (discriminative conditional-mutual-information estimation); **JAFA** Janisch et al (2019) (deep reinforcement learning); **covert2023** Covert et al (2023); two order-learning variants **OL (with/without mask)** Kachuee et al (2019); and **Permutation**, a static globally-ranked acquisition order. All methods use the same mask-aware classifier for prediction and are evaluated under identical availability constraints and panel-level costing. All methods were implemented, trained, and evaluated within the AFABench framework Schütz et al (2026), which provides standardised acquisition policies and a single shared evaluation harness; we extended it with our cohort, the per-patient availability mask, and panel-level costing. We note that standardised comparisons have found discriminative conditional-mutual-information estimators such as DIME Gadgil et al (2024) can outperform generative, EDDI-style estimators on some full-availability benchmarks Schütz et al (2026). We therefore include DIME as a baseline; notably, under our realistic-availability, panel-cost setting it does *not* retain that advantage (Section 3), which is itself consistent with the paper’s central finding. Stochastic-encoding approaches such as SEFA Norcliffe et al (2025) and template-based batch acquisition are further discriminative alternatives we do not evaluate here; adapting each to per-patient availability and marginal panel-level costing is a non-trivial reformulation rather than a drop-in, and we release our harness and cohort so that they can be benchmarked under the same constraints.

We evaluated on the held-out test set across budgets $30–$60, reporting **macro-F1** (the headline metric, weighting all 21 conditions equally), **top-1 accuracy**, and **mean realised cost**. To characterise the achievable range we also measured the mask-aware classifier at three observation levels: demographics and vitals only (0.103 accuracy), the real per-patient availability mask (0.257), and all features observed (0.266); these are properties of the classifier and independent of the cost model. Point estimates pool the five independent patient-level cohort resamples (seeds); we report 95% confidence intervals from patient-level bootstrap resampling for the primary comparison (Table 2) and across-seed standard deviations elsewhere.

**Table 2.**
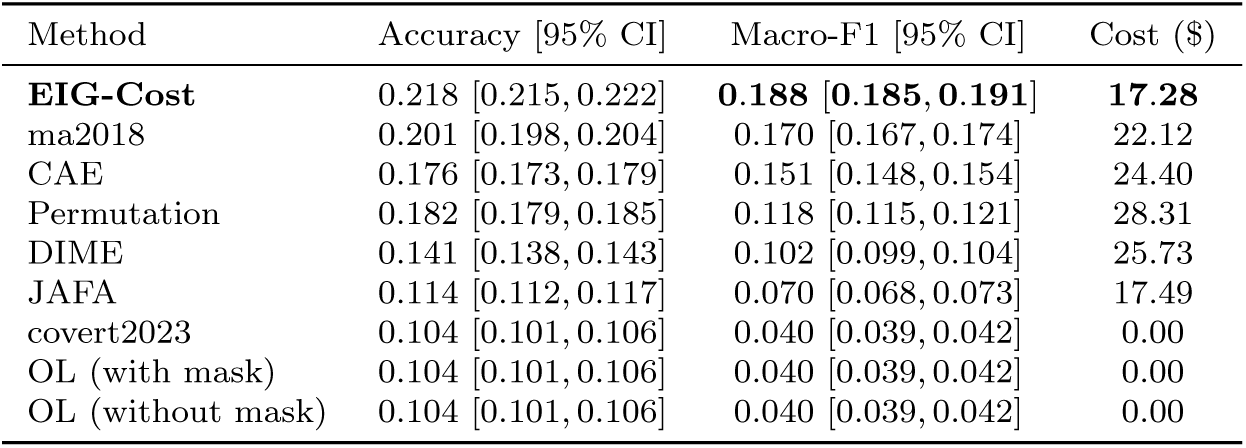
Performance at a $30 budget under panel-level costing. Point estimates pool five patient-level resamples; brackets are 95% confidence intervals from patient-level bootstrap resampling (2,000 replicates). Higher macro-F1 and lower cost are better; best macro-F1 in bold.

## 3 Results

### 3.1 EIG-Cost dominates the cost–performance frontier

Table 2 reports performance at a $30 budget. EIG-Cost attains the highest macro-F1 (0.188) at the lowest realised cost ($17.28), ahead of the strongest baseline, ma2018 (0.170 at $22.12). EIG-Cost exceeds ma2018 in all five resamples; a paired *t*-test on per-seed macro-F1 gives *t* = 8.95, *p <* 0.001 (Cohen’s *d* = 4.0), and a one-sided Wilcoxon signed-rank test gives *p* = 0.031 (the smallest value attainable with five paired samples). The per-patient bootstrap confidence intervals for EIG-Cost and ma2018 do not overlap ([0.185, 0.191] versus [0.167, 0.174]), so the advantage is resolved at the patient level and not an artefact of seed-to-seed variation. DIME, a discriminative conditional-mutual-information method reported to match or beat generative estimators on full-availability benchmarks Gadgil et al (2024); Schütz et al (2026), reaches only 0.102 macro-F1 at $25.73 here, below ma2018, CAE, and even the static permutation order, without collapsing to the vitals-only floor. Its poor showing under availability and panel-cost constraints, despite its strength on conventional benchmarks, is a concrete instance of the paper’s central point: method rankings established under full availability do not transfer to the realistic setting. Figure 3 shows the corresponding cost–performance frontier.

**Fig. 3.**
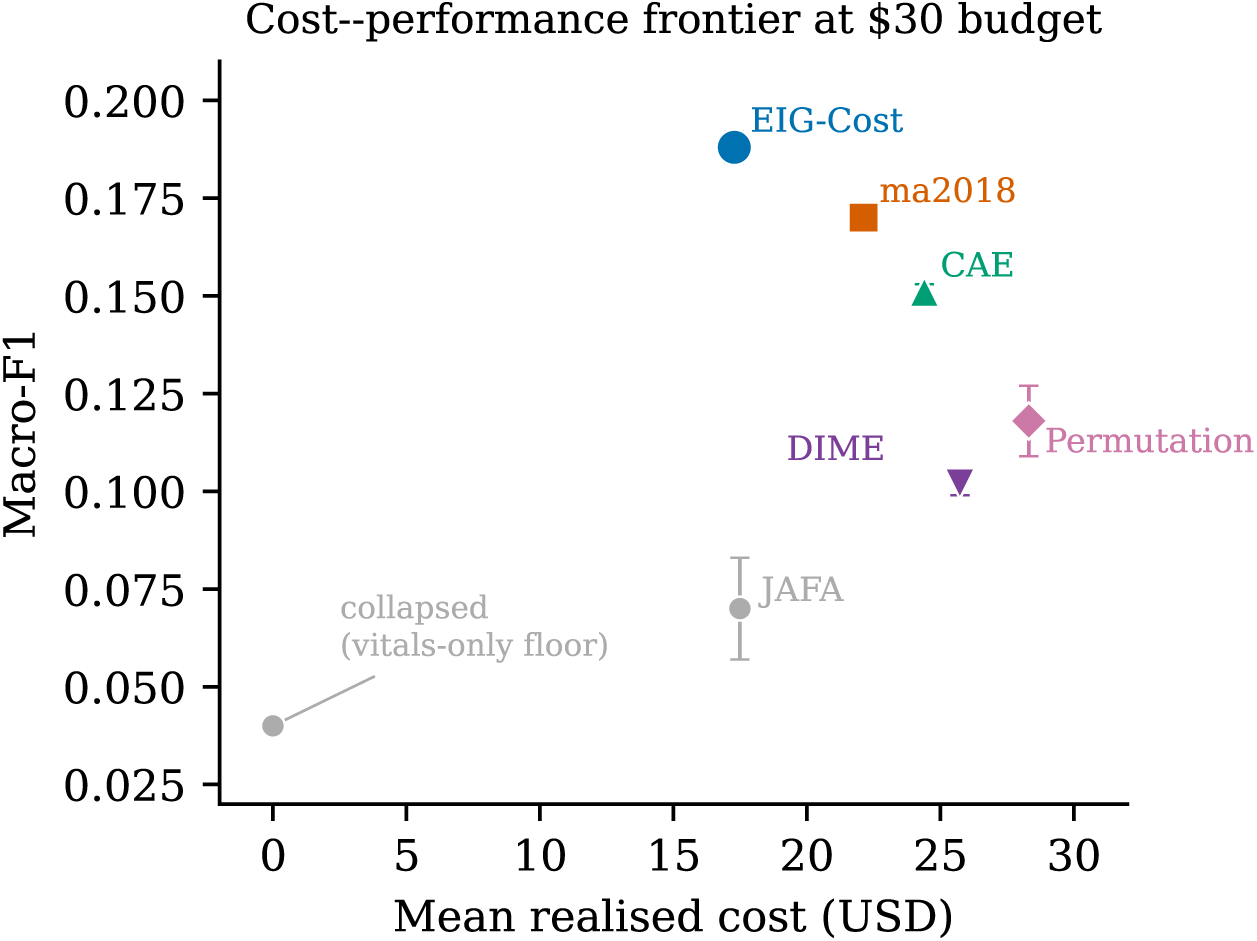
Cost–performance frontier at a $30 budget. EIG-Cost attains the highest macro-F1 at the lowest mean realised cost, defining the upper-left frontier. Three methods collapse to the vitals-only floor (macro-F1 *≈* 0.040) at zero realised cost. Error bars show *±*1 s.d. over five patient-level resamples.

### 3.2 Behaviour across budgets

Table 3 traces macro-F1 across budgets $30–$60. EIG-Cost leads at every budget and reaches each performance level for less money: at $60 it attains 0.231 macro-F1 at a realised cost of $29.16, whereas ma2018 requires $43.47 to reach 0.224. The advantage is largest at the tightest budget ($30: +0.018 macro-F1, roughly four standard deviations) and narrows as the budget grows ($60: +0.007), as competing methods can eventually purchase comparable information (Figure 4). DIME remains the weakest of the adaptive methods at every budget and does not close the gap with more spending: at $60 it reaches only 0.148, still below EIG-Cost’s $30 value of 0.188, so its shortfall reflects the setting rather than an insufficient budget. This profile, a clear advantage precisely where the budget is most constrained, is well suited to cost-constrained decision support.

**Fig. 4.**
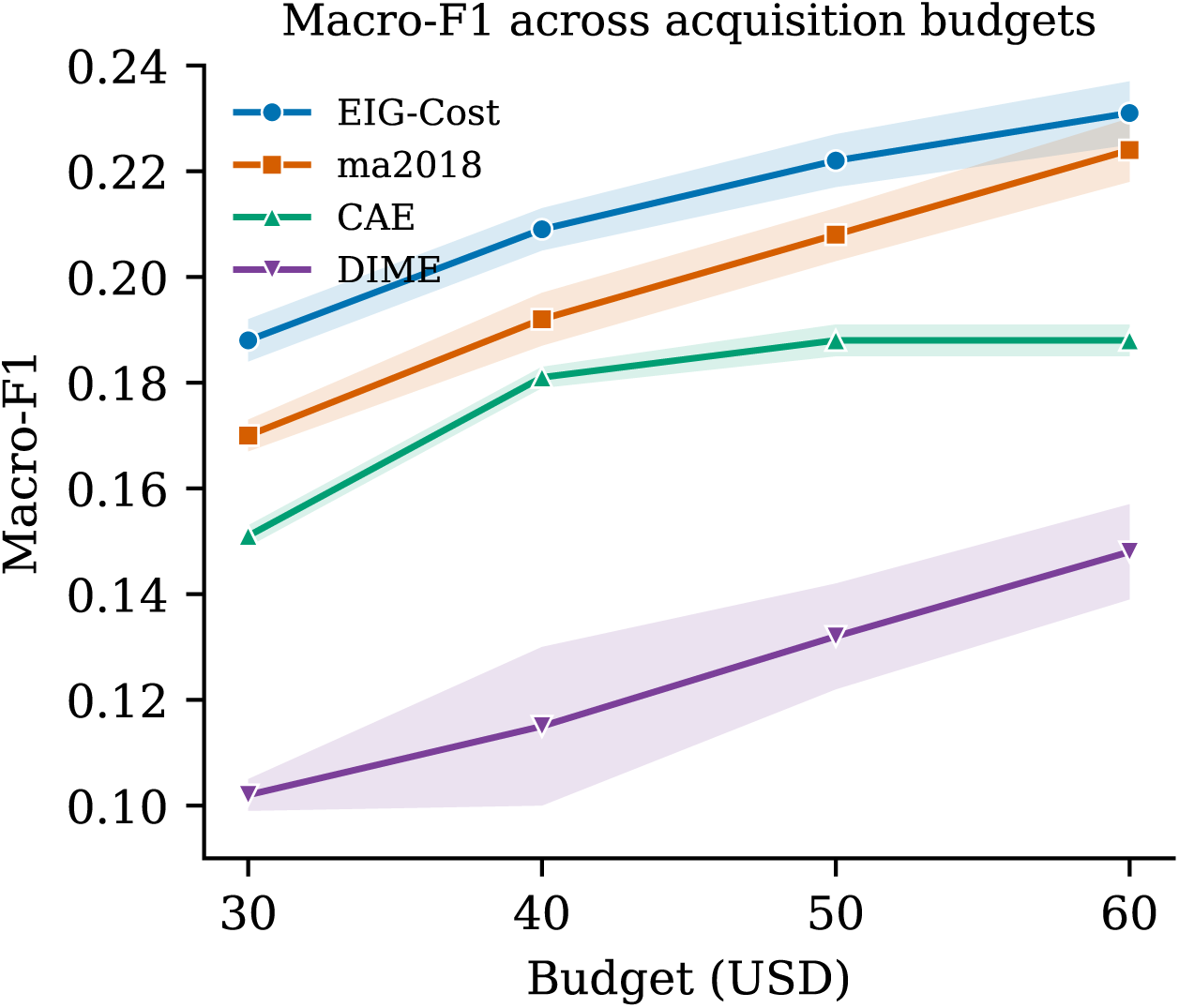
Macro-F1 versus acquisition budget under panel-level costing. Shaded bands show *±*1 s.d. over five resamples. EIG-Cost leads at every budget; the margin is largest at the tightest budget and narrows as competing methods can purchase comparable information. CAE plateaus beyond $40.

**Table 3.** Macro-F1 versus budget (mean *±* s.d. over five resamples), panel-level costing. EIG-Cost leads at every budget; the margin is largest at low budgets.

| Method | \$30 | \$40 | \$50 | \$60 |
| --- | --- | --- | --- | --- |
| **EIG-Cost** | **0.188** | **0.209** | **0.222** | **0.231** |
| ma2018 | 0.170 | 0.192 | 0.208 | 0.224 |
| CAE | 0.151 | 0.181 | 0.188 | 0.188 |
| DIME | 0.102 | 0.115 | 0.132 | 0.148 |

#### Stopping behaviour

At the $30 budget, EIG-Cost stops voluntarily, because no remaining panel has a positive cost-penalised score, in 52.4% of trajectories, while the remaining 47.6% are halted by the budget. It acquires 3.8 panels per patient on average (median 3), but the distribution is skewed: about 40% of patients receive one panel or none, the agent judging the free vitals and demographics already sufficient, while a long tail acquire many. This cost-conscious restraint is appropriate (spending nothing when acquisition would not help is the desired behaviour), but it also contributes to the modest absolute accuracy, since a substantial fraction of patients are classified from baseline information alone. The near-even split between voluntary and budget-limited stopping also explains the earlier insensitivity to *λ* at this budget: for roughly half of patients the budget binds before the cost penalty governs the decision, so the exact value of *λ* has limited influence.

### 3.3 Collapse under realistic availability constraints

Three of the eight methods (covert2023 and both order-learning variants) collapse to the vitals-only floor: they acquire essentially nothing ($0 realised cost) and predict near the class prior, yielding macro-F1 0.040 *±* 0.002. This holds across all five resamples and every budget from $30 to $60. A separate probe that retrained these methods at a $60 budget found them still at the floor, indicating the collapse reflects a genuine failure to adapt to feature availability rather than a budget–training mismatch. Under full-availability assumptions such methods appear competitive; the constraint that exposes their failure is realistic availability, not budget. We report this as an honest finding about evaluation realism rather than a criticism of any individual method, and place the collapsed methods alongside the others in Table 2 for completeness.

### 3.4 Per-condition performance

Because macro-F1 averages over 21 conditions of very different character, we also examined per-condition F1 at the $30 budget (Figure 5, mean over five seeds). Performance spans a wide range and tracks how far each condition’s diagnosis rests on a specific laboratory test that is commonly measured early. The method is strongest on conditions with a decisive, routinely-drawn confirmatory assay: myocardial infarction (F1 0.53; troponin), diabetic emergency (0.45; glucose and related chemistry), acute kidney injury (0.30; creatinine in the basic panel), hyponatraemia (0.29; sodium), and pancreatitis (0.27; lipase). It is weakest on two high-prevalence, high-acuity conditions (pneumonia, F1 0.001, and sepsis, 0.011) that are diagnosed largely from imaging, culture, and the overall clinical picture rather than a single early chemistry value, and whose more specific laboratory markers (procalcitonin, blood culture, lactate) are among the least available within the 12-hour window. For these two conditions the misclassifications do not concentrate on a single look-alike diagnosis but disperse across several conditions whose discriminative labs were more often available (most frequently pancreatitis, urinary tract infection, atrial fibrillation, and chronic obstructive pulmonary disease), consistent with the model having little availability-constrained signal with which to identify them rather than systematically confusing them with one specific alternative. This per-condition structure is itself evidence that the binding constraint on the hard classes is the availability of discriminative tests at the decision point, not model capacity, and it is these collapses (not uniform mediocrity) that depress the macro-average, which is why we report macro-F1 as the honest headline rather than accuracy.

**Fig. 5.**
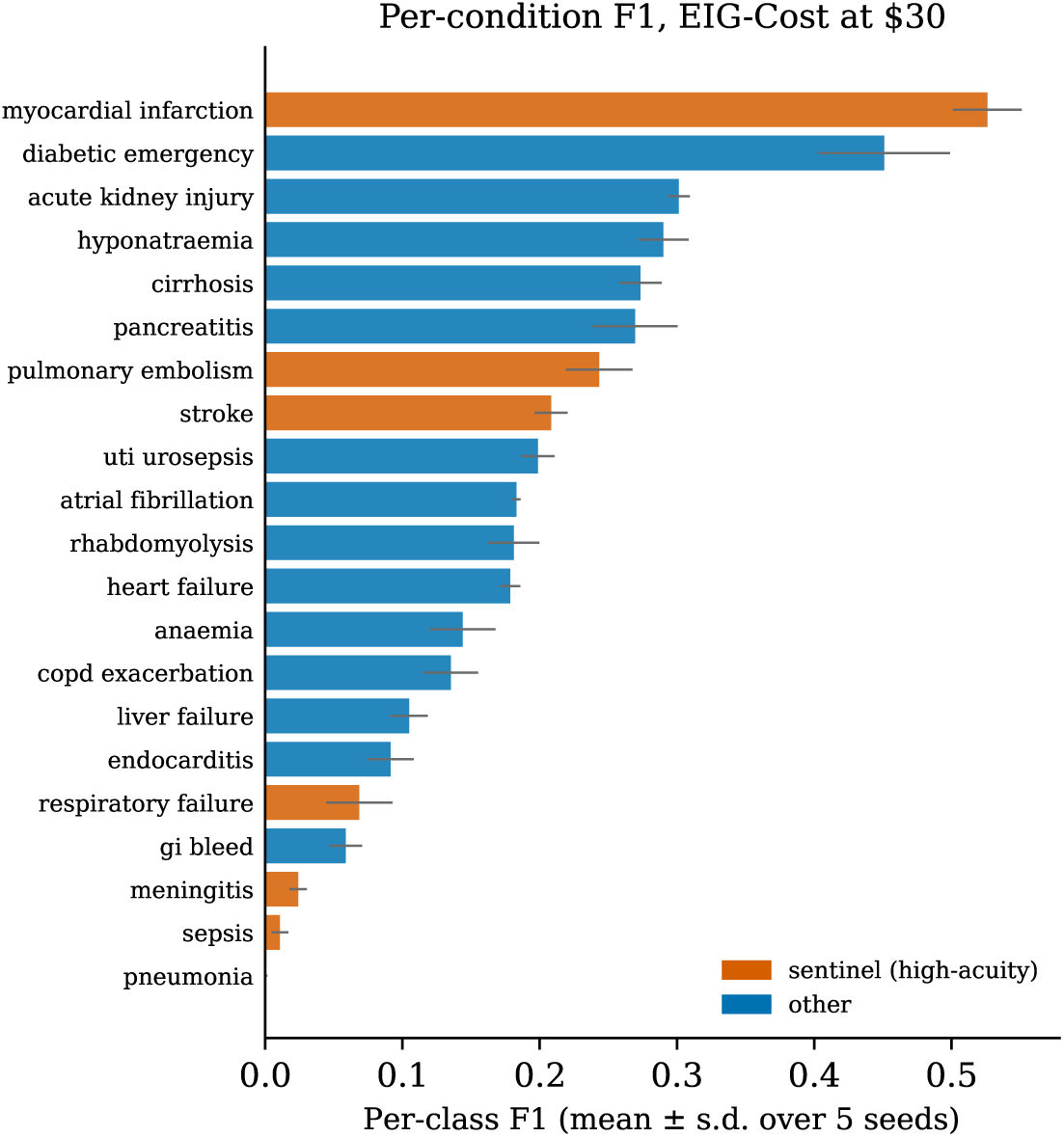
Per-condition F1 for EIG-Cost at a $30 budget (mean *±* s.d. over five seeds), ranked. Sentinel (high-acuity) conditions are highlighted. Performance is highest where a specific, commonly-available early laboratory test is decisive (e.g. troponin for myocardial infarction) and lowest for pneumonia and sepsis, whose confirmatory investigations are largely imaging-, culture-, or availability-limited within the decision window.

### 3.5 Sensitivity to hyperparameters

EIG-Cost has two free hyperparameters evaluated at inference: the cost penalty *λ* and the number of Monte-Carlo completions *K*. We swept each at the $30 budget across five seeds (Table 4, Figure 6). Results are effectively flat: over a tenfold range of *λ* (0.002–0.02) macro-F1 moves from 0.186 to 0.188, within one standard deviation, and mean cost stays near $17.3; over *K* ∈ {20, 50, 100} macro-F1 holds at 0.187–0.188. The insensitivity to *λ* is expected at tight budgets: when the budget rather than the stopping rule is the binding constraint, the agent rarely reaches the point where the cost penalty determines whether to stop, so the exact penalty matters little; *λ* would exert more influence at looser budgets. The insensitivity to *K* indicates that 50 completions already give a stable entropy estimate. In short, the reported results do not depend on fine-tuning these values.

**Fig. 6.**
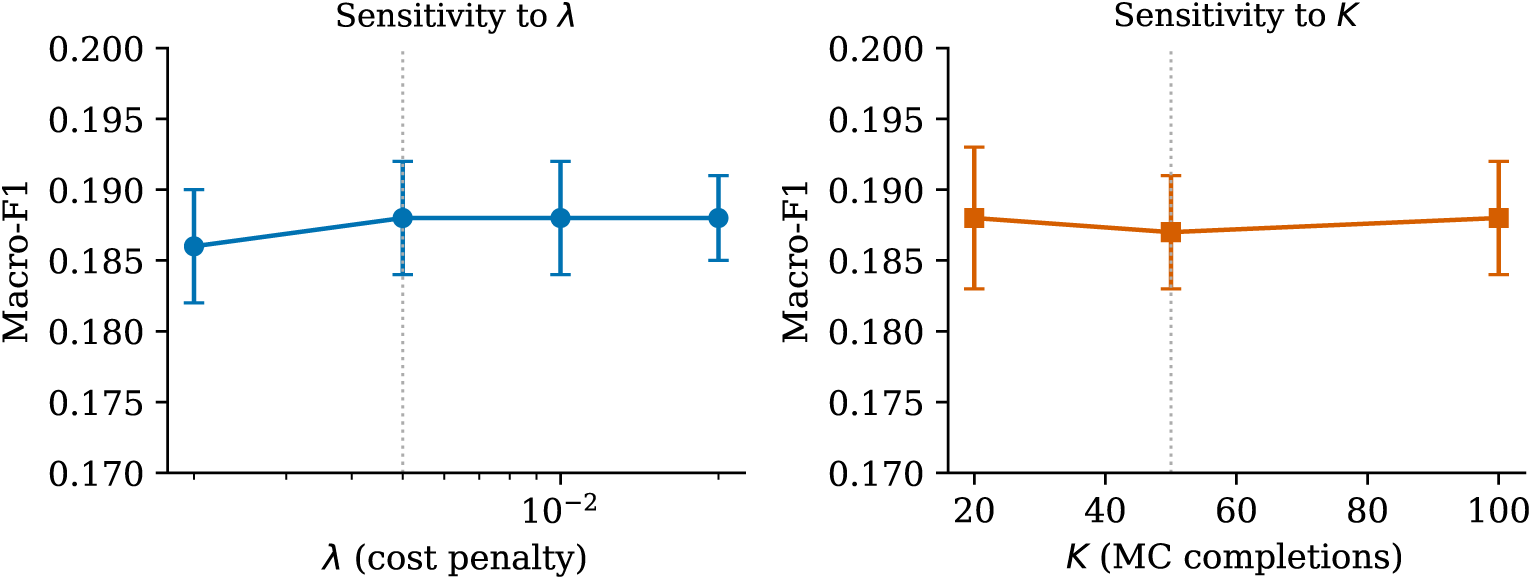
Macro-F1 versus the cost penalty *λ* (left) and the number of Monte-Carlo completions *K* (right), at the $30 budget (mean *±* s.d. over five seeds). Dotted lines mark the default values. Performance is flat within error across both sweeps.

**Table 4.** Sensitivity of EIG-Cost to the cost penalty *λ* and the number of Monte-Carlo completions *K*, at the $30 budget (mean *±* s.d. over five seeds). Default values (*λ* = 0.005, *K* = 50) in bold.

| Sweep | Value | Macro-F1 | Mean cost (\$) |
| --- | --- | --- | --- |
| $\lambda$ | 0.002 | $0.186 \pm 0.004$ | 17.17 |
|  | **0.005** | **$0.188 \pm 0.004$** | **17.29** |
| | 0.010 | $0.188 \pm 0.004$ | 17.35 |
| | 0.020 | $0.188 \pm 0.003$ | 17.35 |
| $K$ | 20 | $0.188 \pm 0.005$ | 17.29 |
|  | **50** | **$0.187 \pm 0.004$** | **17.29** |
| | 100 | $0.188 \pm 0.004$ | 17.29 |

### 3.6 Calibration

Beyond discrimination, a decision-support tool’s predicted probabilities are useful only if they are calibrated: if a stated confidence of *p* corresponds to being correct about a fraction *p* of the time. We assessed top-label calibration of the sequential predictions at the $30 budget, pooling the five seeds (Figure 7). The expected calibration error is 0.048 and the top-label Brier score is 0.149. The reliability curve is monotonic and close to the diagonal: empirical accuracy rises with confidence across the range, and high-confidence predictions are dependable (predictions made with confidence above 0.9 are correct 84% of the time). Overall the model is marginally under-confident (mean confidence 0.194 versus accuracy 0.205), with the largest deviation in the mid-confidence range; under-confidence is the more benign failure mode for clinical use. Despite the modest absolute accuracy imposed by the realistic-availability setting, then, the probabilities EIG-Cost reports are well-calibrated and can be taken at face value; most usefully, its confident predictions can be trusted.

**Fig. 7.**
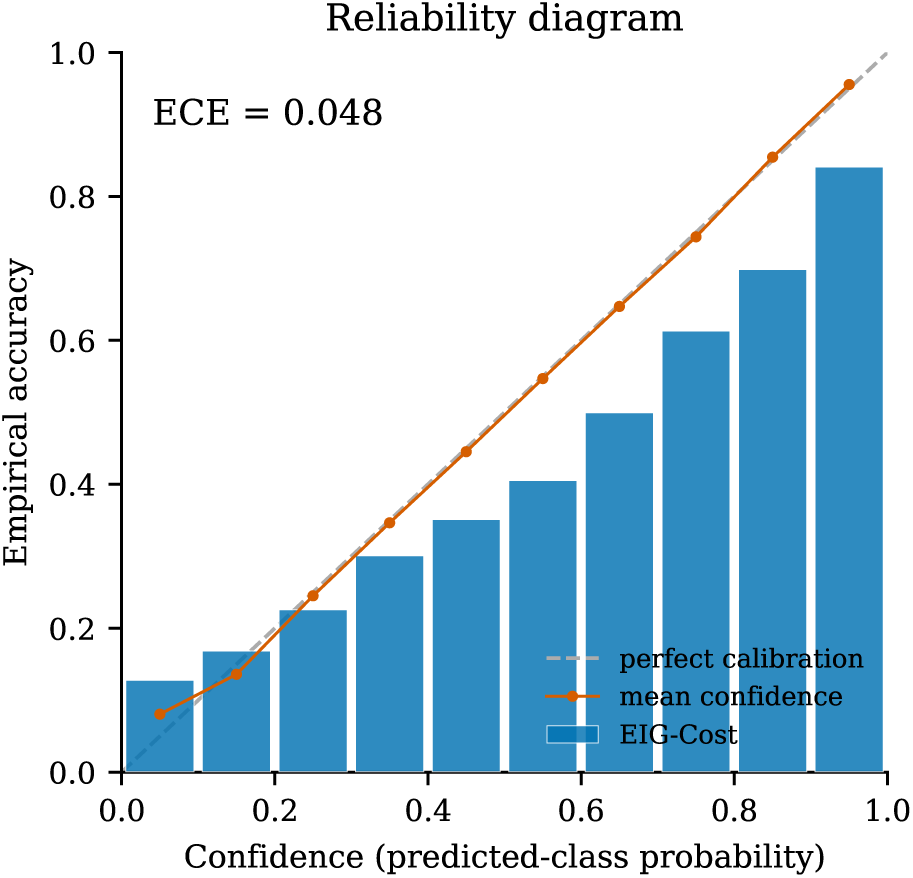
Reliability diagram for EIG-Cost at a $30 budget (top-label calibration, pooled over five seeds). Bars show empirical accuracy per confidence bin; the orange line is mean confidence and the dashed line is perfect calibration. The expected calibration error is 0.048.

### 3.7 Subgroup performance

As an exploratory fairness assessment, we examined performance across subgroups defined by sex and age at the $30 budget, pooling the five seeds (Figure 8; 95% confidence intervals from patient-level bootstrap resampling). We emphasise that these strata were not pre-specified and the analysis is exploratory. Performance was essentially identical by sex (macro-F1 0.184 for both female and male patients, with overlapping confidence intervals). Across age bands, macro-F1 declined modestly and monotonically with age, from 0.189 (*<* 45 years) to 0.144 (*≥* 80), a difference whose confidence intervals separate the youngest and oldest bands. This gradient is most plausibly attributable to case mix rather than a demographic bias in the model: older admissions are enriched for exactly the conditions the per-condition analysis identified as hardest under realistic availability (heart failure, pneumonia, and sepsis), so the drop tracks disease difficulty rather than age *per se*. We report the pattern here as a signal to monitor in prospective evaluation rather than as an established bias.

**Fig. 8.**
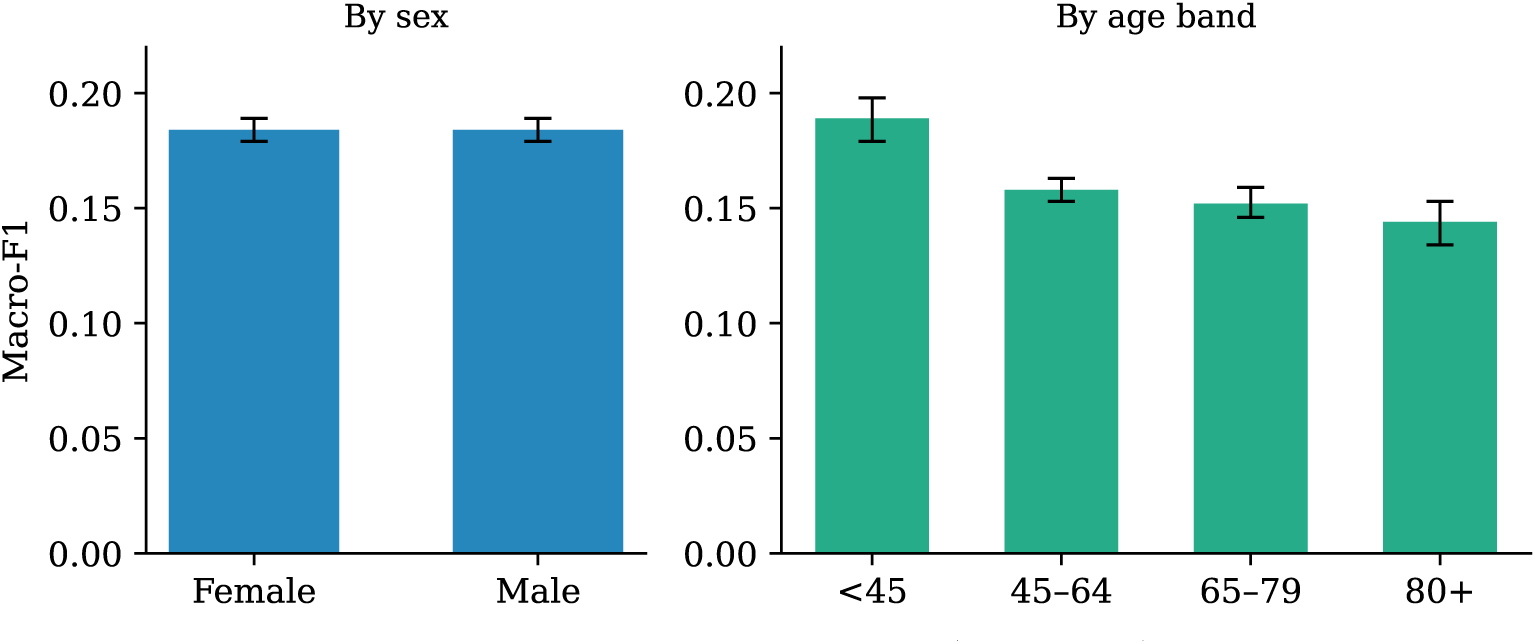
Exploratory subgroup performance of EIG-Cost at a $30 budget (macro-F1, mean over five seeds; error bars are 95% patient-level bootstrap confidence intervals). Performance is equal by sex and declines modestly with age, consistent with older admissions being enriched for the harder-to-diagnose conditions.

Calibration was likewise consistent across subgroups. Expected calibration error was 0.049 for female and 0.048 for male patients (matching the overall 0.048), and lay between 0.039 and 0.058 across the three older age bands. The one exception was the youngest band (*<* 45 years), which was both the smallest (*n ≈* 1,600) and mildly under-confident (ECE 0.087; mean confidence 0.22 against accuracy 0.28), meaning the model slightly understated its own accuracy for these patients. Under-confidence is the more benign direction of miscalibration, and the elevated error is partly a small-sample effect.

## 4 Discussion

Two messages emerge. First, *evaluation realism materially changes conclusions*. Enforcing patient-level separation, a decision-time cutoff, and empirical availability lowers headline performance and reveals that some methods which appear competitive under full-availability assumptions in fact fail to adapt to what is actually measurable. The narrow gap between the availability-limited classifier (0.257 accuracy) and the all-features ceiling (0.266) shows how little of the full panel is realised within the decision window, and hence how much apparent performance in prior benchmarks depended on information that would not be available at the point of decision. This is a caution for benchmarking practice more than a verdict on any single algorithm.

Second, in this harder regime, *cost-aware information-gain scoring is a robust default*. EIG-Cost attains the best macro-F1 at the lowest realised cost across the budget range, and its advantage is largest at tight budgets, exactly where a decision-support tool intended to curb overtesting would operate. Because the acquisition score penalises expected information gain by real dollar cost and respects per-patient availability, the method naturally avoids spending on unavailable or low-yield panels.

### Limitations

Several limitations bound the scope of these findings. *Selection on the measured regime.* The availability mask encodes which tests were historically ordered, not which *could* have been ordered at the decision point. Conditioning acquisition on this mask means the evaluation reflects the historical test-ordering regime and cannot credit a test that a proactive system would order but that was never measured; positivity does not hold for such never-measured tests. This is a deliberate trade-off (it is what makes the setting realistic rather than counterfactual), but it also means the generative model and the learned policies may be biased toward commonly obtained tests, with fairness implications for patients whose care deviates from historical norms. We regard characterising and correcting this selection-on-treatment bias (for example with inverse-probability weighting or semi-synthetic counterfactual availability) as important future work rather than something the present design resolves. *No turnaround times.* We model monetary cost but not time-to-result; acquired tests are revealed immediately, whereas real laboratories have non-trivial and heterogeneous turnaround times that a deployed system should weigh. Extending the cost term to a delay-aware penalty is a natural direction. *Per-feature scoring.* As noted in Methods, EIG is scored per feature with panel-level billing rather than as a joint panel acquisition; this does not capture intra-panel complementarity, and a joint-acquisition formulation may further improve panels whose components are only jointly informative. *Cost schedule.* Costs use the 2026 Medicare schedule; absolute costs differ across payers and institutions, though the acquisition ranking is driven primarily by information content rather than absolute price. *Single database and fixed architecture.* Results are from MIMIC-IV with one classifier and one generative architecture; external validation on an independent intensive-care cohort, and robustness across alternative classifier and imputation architectures and masking rates, remain untested. *Cost-model mismatch for baselines.* Budget-dependent baselines are trained at one budget and evaluated under panel-level costing, a train/evaluation cost-model mismatch we note explicitly; the collapse we report may be partly attributable to insufficient adaptation to the availability constraint rather than to the methods’ intrinsic ceilings, and availability-aware retraining of those baselines could raise them. The per-condition, calibration, and exploratory subgroup analyses reported above partly address a fuller clinical-prediction-model evaluation; prospective, pre-specified subgroup and fairness analysis on external data remains important future work.

## 5 Conclusions

Under a realistic-constraint protocol for clinical AFA (patient-level splitting, a decision-time cutoff, empirical per-patient availability, and panel-level dollar costing) cost-aware Expected Information Gain scoring dominates the cost–performance frontier, while several published methods collapse to a vitals-only baseline that higher budgets do not rescue. Clinical AFA is harder than prior full-availability benchmarks imply, and cost-awareness is most valuable precisely under the harder, more realistic setting.

## Appendix A Condition prevalence

Table A1 gives the exact prevalence of each of the 21 diagnostic conditions in the pooled test set, ranging from heart failure (11.6%) to meningitis (0.3%). The marked imbalance is the reason macro-F1, which weights all conditions equally, is used as the headline metric.

**Table A1.** Prevalence of the 21 conditions in the pooled test set (*n* = 64,962), in descending order.

| Condition | % | Condition | % |
| --- | --- | --- | --- |
| Heart failure | 11.6 | COPD exacerbation | 3.7 |
| Pneumonia | 10.2 | Pulmonary embolism | 3.3 |
| Myocardial infarction | 9.9 | Cirrhosis | 2.9 |
| Acute kidney injury | 8.7 | Respiratory failure | 2.2 |
| UTI / urosepsis | 8.2 | Diabetic emergency | 1.9 |
| Sepsis | 7.6 | Liver failure | 1.8 |
| Atrial fibrillation | 7.4 | Hyponatraemia | 1.6 |
| Stroke | 6.6 | Anaemia | 1.5 |
| Pancreatitis | 5.7 | Endocarditis | 0.5 |
| GI bleed | 4.3 | Rhabdomyolysis | 0.4 |
|  |  | Meningitis | 0.3 |

## Appendix B Model card

### Model details

EIG-Cost is a cost-aware active feature acquisition policy for early differential diagnosis. It couples a mask-aware multilayer perceptron classifier (three layers, 256/128/64 units, batch normalisation, ReLU, dropout 0.2) with a conditional variational autoencoder for imputation, and selects features by Monte-Carlo Expected Information Gain penalised by marginal panel-level dollar cost (penalty *λ* = 0.005, *K* = 50 completions). Developed in this study; not versioned for release beyond the accompanying code.

### Intended use

Research and methodological benchmarking of cost-aware test-ordering under realistic availability constraints. It is *not* a deployable diagnostic device, is not cleared by any regulator, and must not be used to guide patient care. Intended users are informatics and machine-learning researchers.

### Data

MIMIC-IV v2.2 (Beth Israel Deaconess Medical Center intensive care). Cohort: 64,766 acute admissions from 39,884 unique patients, 21 discharge-diagnosis condition classes, 55 features grouped into 30 billable panels. Inputs are the first 12 hours of an admission; labels are the primary ICD-9/10 discharge diagnosis mapped to the 21 classes. Splits are patient-level (train/validation/test) with zero subject overlap, repeated over five resampling seeds.

### Evaluation factors

Performance is reported overall, per condition, by acquisition budget ($30–$60), and across subgroups defined by sex and age band. Calibration is assessed overall and by subgroup.

### Metrics

Macro-F1 (primary, chosen for class imbalance), accuracy, realised dollar cost, expected calibration error, and Brier score, each with across-seed variability and, for the primary comparison, patient-level bootstrap 95% confidence intervals.

### Performance summary

At the $30 budget EIG-Cost attains macro-F1 0.188 (95% CI [0.185, 0.191]) at $17.28 mean cost, the best cost-performance trade-off among nine methods. Per-condition F1 ranges from 0.53 (myocardial infarction) to near zero for pneumonia and sepsis, tracking the availability of each condition’s discriminative test within 12 hours. Expected calibration error is 0.048 overall, 0.048–0.049 by sex, and 0.039–0.087 across age bands. Inference runs on CPU at roughly 180 ms per patient.

### Limitations

Single database and single architecture; discharge-diagnosis labels; a per-patient availability mask that reflects historical ordering (selection on the measured regime, with positivity not holding for never-measured tests); no modelling of test turnaround time; per-feature rather than joint-panel information scoring; and a US-Medicare cost schedule. Absolute accuracy is modest, and performance is weakest for imaging- or culture-dependent diagnoses.

### Ethical considerations and monitoring

Because the availability mask encodes historical clinician behaviour, a deployed successor could systematically disadvantage patients whose care deviated from historical norms; subgroup performance and calibration by age, sex, and other protected attributes would need prospective monitoring, alongside drift in cost schedules and measurement practices. No protected-attribute information is used as a model input.

## Appendix C TRIPOD+AI checklist

Table C2 maps the TRIPOD+AI reporting items to where each is addressed. Items that cannot be satisfied by a retrospective, unregistered study are marked and justified rather than omitted.

## Funding

This work was supported by the Israel Science Foundation (ISF) and the Technion – Israel Institute of Technology.

## Data availability

This study used MIMIC-IV, available to credentialed users under the PhysioNet Credentialed Health Data Use Agreement. The data are not redistributed by the authors and must be obtained from PhysioNet.

## Code availability

The EIG-Cost implementation is available at https://github. com/JosephBingham/EIG Cost.

## Competing interests

The authors declare no competing interests.

## Author contributions

J.B. designed the study, implemented the methods and experiments, and wrote the manuscript. N.A. contributed to the experimental design and analysis. All authors reviewed and approved the final manuscript.

## Ethics

MIMIC-IV is a de-identified, publicly available database; its collection was approved by the institutional review boards of the Beth Israel Deaconess Medical Center and the Massachusetts Institute of Technology. Use in this study is governed by the PhysioNet Credentialed Health Data Use Agreement.

## Declarations

Clinical trial number: not applicable Consent to Participate declaration: not applicable

---

**Table C2.**
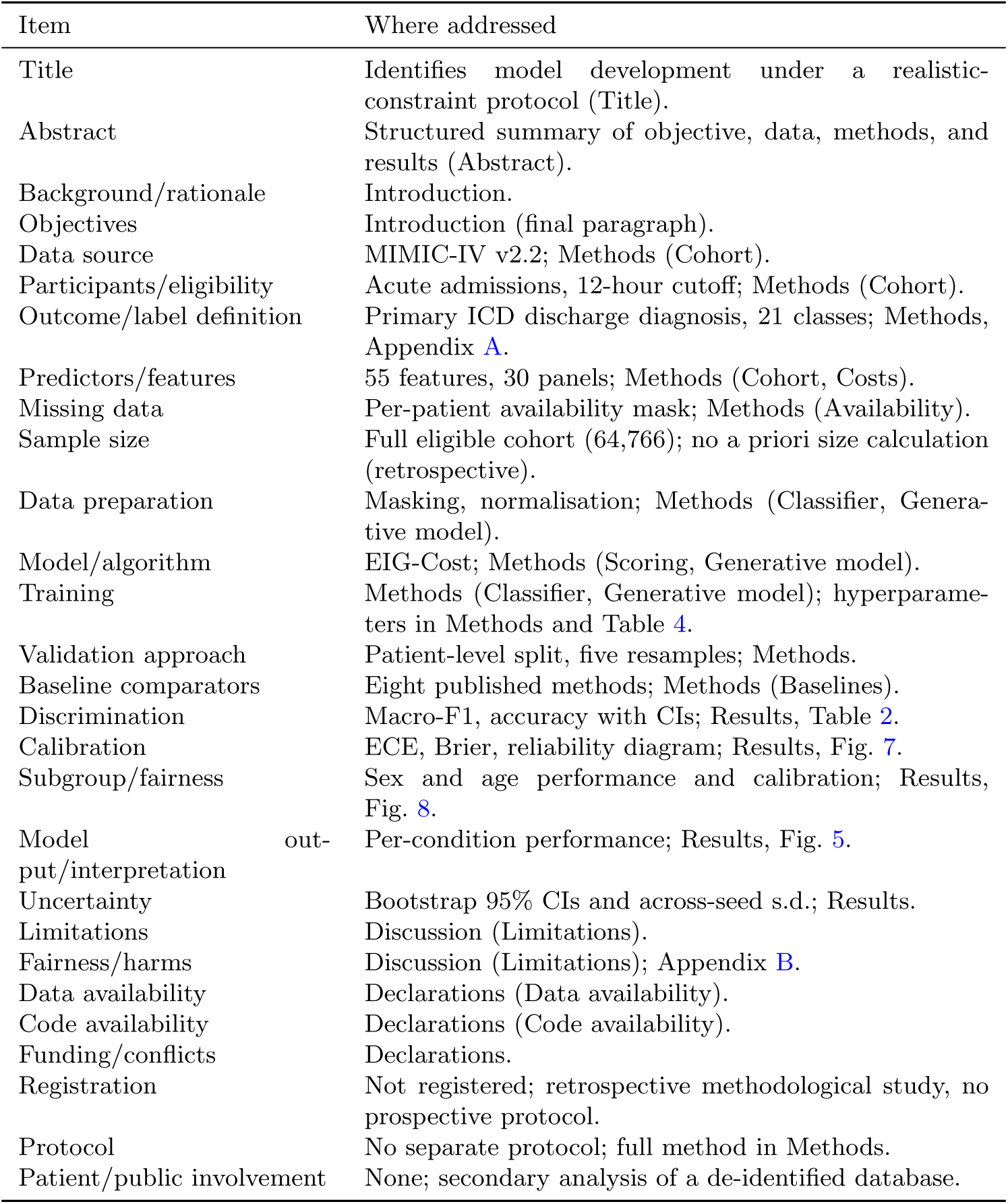
TRIPOD+AI reporting checklist, condensed, with location in this article. D/V = development/validation.

## Notes

### Competing Interest Statement

The authors have declared no competing interest.

### Author Declarations

That data is from a publicly available dataset that was created and published before this study

